# HybridNet-XR: Efficient Teacher-Free Self-Supervised Learning for Autonomous Medical Diagnostic Systems in Resource-Constrained Environments

**DOI:** 10.64898/2026.03.16.26348570

**Authors:** Simeon Mayala, Deogratias Mzurikwao, Emmanuel Suluba

## Abstract

Deep learning model classification on large datasets is often limited in countries with restricted computational resources. While transfer learning can offset these limitations, standard architectures often maintain a high memory footprint. This study introduces HybridNet-XR, a memory-efficient and computationally lightweight hybrid convolutional neural network (CNN) designed to bridge the domain gap in medical radiography using autonomous self-supervised learning protocols. The HybridNet-XR architecture integrates depthwise separable convolutions for parameter reduction, residual connections for gradient stability, and aggressive early downsampling to minimize the video RAM (VRAM) footprint. We evaluated several training paradigms, including teacher-free self-supervised learning (SSL-SimCLR), teacher-led knowledge distillation (KD), and domain-gap (DG) adaptation. Each variant was pre-trained on ImageNet-1k subsets and fine-tuned on the ChestX6 multi-class dataset. Model interpretability was validated through gradient-weighted class activation mapping (Grad-CAM). The performance frontier analysis identified the HybridNet-XR-150-PW (Pre-warmed) as the optimal configuration, achieving a 93.38% average accuracy and 99% AUC while utilizing only 814.80 MB of VRAM. Regarding class-wise accuracy, this variant significantly outperformed standard MobileNetV2 and teacher-led models in critical diagnostic categories, notably Covid-19 (97.98%) and Emphysema (96.80%). Grad-CAM visualizations confirmed that the teacher-free pre-warming phase allows the model to develop sharper, anatomically grounded focus on pathological landmarks compared to distilled models. Specialized pre-warming schedules offer a viable, computationally autonomous alternative to knowledge distillation for medical imaging. By eliminating the requirement for high-performance teacher models, HybridNet-XR provides a robust and trustworthy diagnostic foundation suitable for clinical deployment in resource-constrained environments.

**Author summary:** Traditional deep learning models for medical imaging are often too large for the low-power computers available in many global health settings. We developed a new model to bridge this computational gap. We designed HybridNet-XR, a highly efficient AI architecture, and trained it using a “teacher-free” method that doesn’t require a massive supercomputer. We found a specific version (*H*-*XR*_150_-*PW*) that provides high accuracy while using very little memory. Our results show that high-performance diagnostic AI can be deployed on standard, low-cost hardware. Furthermore, using visual heatmaps (Grad-CAM), we proved that the AI correctly identifies medical landmarks like lung opacities, ensuring it is safe and reliable for real-world clinical use.

## Introduction

The classification of images using deep learning models from huge datasets is a limitation, especially in countries with limited computing resources. In this group of countries, the challenge remains active. Then, it is ideal to work with computational efficient models whose capacity limitation can be offset by transfer learning.

Xception is a deep convolutional neural network (CNN) that was developed by François Chollet. Its core innovation lies in the extensive use of depthwise separable convolutions, depthwise convolution and a pointwise convolution. This approach significantly reduces the number of parameters and computational cost, making the model highly efficient without compromising performance. Also, the architecture incorporates residual connections to aid in training deep networks. It has demonstrated superior performance on large-scale datasets and is widely used for various computer vision tasks, including image classification and object detection [1].

ResNet-50 is a CNN that is 50 layers deep. Its main innovation is the use of residual connections which are also known as shortcut connections. These connections allow the network to skip over certain layers and add the output of a previous layer to a later one. This mechanism helps to mitigate the vanishing gradient problem, enabling the network to learn and achieve high accuracy. It is primarily used for image classification and is often a foundational model for other computer vision tasks like object detection [2].

MobileNetV2 is a highly efficient convolutional neural network architecture optimized for mobile and embedded devices. Its core innovations are the inverted residual block and linear bottlenecks. The inverted residual block first expands the input feature channels, applies a computationally cheap depthwise convolution, and then projects the features back to a low-dimensional space. To avoid information loss, the final projection layer uses a linear bottleneck, omitting non-linear activation functions. This design allows the model to achieve high accuracy with significantly fewer parameters and lower computational costs, making it ideal for on-device computer vision tasks like image classification and object detection [3].

The model MobileNetV3 presented in paper [4] was developed for mobile CPUs by optimizing the accuracy-latency trade-off. The approach combines automated hardware-aware Neural Architecture Search (NAS) for global structure with the NetAdapt algorithm for layer-wise fine-tuning, while also introducing manual design innovations like the “h-swish” activation function and streamlined network headers. The model demonstrates significant improvements over its predecessor.

EfficientNet [5] presents work that aimed to develop a principled method for scaling convolutional neural networks (ConvNets) to achieve better accuracy and efficiency. They proposed a compound scaling method that uniformly scales a baseline network’s depth, width, and resolution using a single compound coefficient (*ϕ*). They first used a multi-objective neural architecture search to develop a highly efficient baseline network called EfficientNet-B0. By applying compound scaling to this baseline, they created a family of models (EfficientNet-B1 to B7) that consistently outperform existing models. The models demonstrated strong transferability, reaching state-of-the-art accuracy on common transfer learning datasets.

The ShuffleNet work presented in [6] was aimed at designing a highly computation-efficient CNN architecture for mobile platforms with extremely limited power. The authors propose pointwise group convolutions and channel shuffle, which overcomes the information-blocking side effects of group convolutions by enabling cross-group information flow. The results obtained demonstrate a significant performance on ImageNet. The propose architecture delivers an impressive speedup while maintaining comparable accuracy.

While AI offers transformative potential for diagnostics, high-performance models must be adaptable to cost-effective, resource-efficient solutions to promote healthcare equity in low-resource and remote settings [7].

In this study, to address the challenge of classifying images from a huge dataset with limited computational resources, we present a CNN model that shares design principles with prominent modern architectures like Xception, ResNet50, and MobileNetV2. The main idea of the proposed model is to be a memory-efficient and computationally lightweight hybrid CNN. We adopt Xception’s use of separable convolutions to reduce parameters and combine it with ResNet50’s idea of residual connections to mitigate the vanishing gradient problem. We also modify some structures to simplify the architecture and make it less complex than its counterparts. The design is aimed at directly tackling the memory footprint, allowing for efficient training and deployment. We hypothesize that the fusion of these ideas will create a model that is both efficient and effective for this specific task.

The proposed model focuses on replacing the standard convolutions with the more efficient depthwise separable version while keeping the skip connections mostly standard. While the MobileNetV2 uses inverted Residuals and Linear bottlenecks to minimize the memory access and maximize representational power in a limited parameter budget, the proposed hybrid model (HybridNet-XR) makes use of extensive depthwise separable convolutions the fact that makes it computationally efficient plus aggressive layer count reduction (shrinking the overall structure) and early downsampling compared to its Xception inspiration.

## Materials and methods

### Material

We use ImageNet-1k dataset (also known as ILSVRC) to train the proposed model and the selected deep learning architectures for comparison. The dataset contains 1.28 million training images, 50,000 validation images, and 100,000 test images. The ImageNet-1k dataset consist of 1000 classes of distinct object categories [8].

We also use National Institutes of Health (NIH) Chest X-Ray dataset comprising of 112,120 X-ray images with disease labels from 30,805 unique patients. The data is freely available and well known as NIH Chest X-ray 14. The images used are presented in JPG format with a resolution of 224 × 224 pixels (resized) as stated from the source [9]. In this study we use this data for training the variants of the developed model for reducing the domain gap.

We use ChestX6: Multi-Class X-ray dataset during the transfer learning of each pre-trained variant model. It is the Chest X-Ray dataset intended for Multi-Class Pulmonary disease classification consisting of six classes: Normal, Pneumonia-Bacterial, Pneumonia-Viral, COVID-19, Tuberculosis and Emphysema. It includes a total of 18,036 grayscale Chest X-ray images, resized to 224 × 224 pixels and presented in .JPG format [10].

## Method

### Mathematical Formulation of the HybridNet-XR Architecture

The HybridNet-XR architecture is built upon three foundational pillars designed to optimize representational capacity within resource-constrained environments.

### Pillar 1: Parameter and Computational Cost Reduction

The architecture transitions from standard convolutions to Depthwise Separable Convolutions (DSC) [11] to achieve efficiency. The computational reduction factor, ℛ, is defined by the ratio of standard Multiply-Accumulate (MAC) operations to DSC operations:

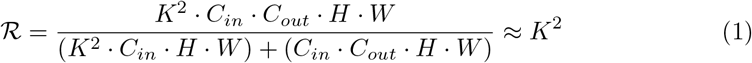

Where *K* is the kernel size, *C*_*in*_ and *C*_*out*_ are channels, and *H* × *W* are spatial dimensions. By utilizing this decomposition, the model significantly reduces parameters without sacrificing feature depth.

### Pillar 2: Mitigation of the Vanishing Gradient Problem

To maintain gradient stability during deep training, we implement additive residual connections. The transformation through a specific residual block *l* within the set of total layers *l* ∈ {1, …, *L*_*total*_} is formulated as:

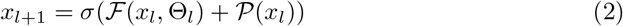

Where:

- *l* denotes the index of the current convolutional block.
- *x*_*l*_ and *x*_*l*+1_ are the input and output feature maps of the *l*-th block respectively.
- ℱ represents the non-linear mapping through Depthwise Separable Convolution (DSC) layers.
- *σ* is the ReLU activation function.
- 𝒫 is the 1 × 1 projection shortcut used for dimensional alignment when spatial or channel dimensions change.
- Θ_*l*_ represents the learnable parameters specific to block *l*.

This mechanism ensures robust gradient propagation 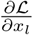 throughout the network by providing a direct path for the gradient to flow to earlier layers.

### Pillar 3: Memory and Spatial Optimization

To minimize the Video RAM (VRAM) footprint required for backpropagation, the model enforces aggressive early downsampling. Total activation memory consumption *M* is defined as:

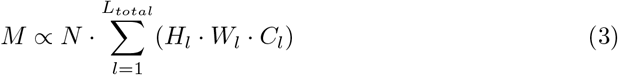

Where *N* is the batch size, *L*_*total*_ is the layer count, and *H*_*l*_, *W*_*l*_, *C*_*l*_ are the spatial and channel dimensions at layer *l*. Utilizing strides (*s* = 2) early in the forward pass halves the spatial dimensions, resulting in an exponential decrease in stored activations.

### Global Feature Aggregation and Flow

The architectural flow transforms input *X* into a probability distribution 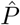 over the set of target classes *L*_*classes*_:

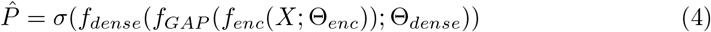

The final predicted label *ŷ*_*n*_ for the *n*-th sample is determined by:

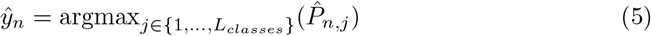

The individual components are defined as follows:

## Nomenclature

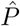 The model output, representing the probability distribution over *L*_*classes*_ target classes.

*X* Input tensor representing a batch of images with shape ℝ ^*N* ×*H*×*W* ×*C*^.

*f*_*enc*_ Convolutional encoder backbone utilizing the three-pillar strategy to extract feature maps 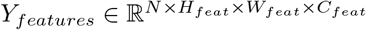 with parameters Θ_*enc*_.

*f*_*GAP*_ Global Average Pooling operation collapsing (*H*_*feat*_ × *W*_*feat*_) into a rank-2 tensor 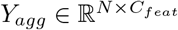.

*f*_*dense*_ Fully connected layer mapping aggregated features to logits 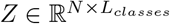 with parameters Θ_*dense*_.

Θ Complete set of learnable parameters Θ = {Θ_*enc*_, Θ_*dense*_}.

*σ* Softmax activation function ensuring 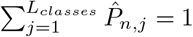.

### Loss Function Formulation

The training of the HybridNet-XR series utilizes three distinct loss components to govern feature discovery, knowledge transfer, and domain alignment.

### Contrastive Objective (NT-Xent)

For all variants, the primary self-supervised objective is the Normalized Temperature-scaled Cross Entropy loss [12]. For a pair of augmented views (*z*_*i*_, *z*_*j*_) in a batch of size 2*N* :

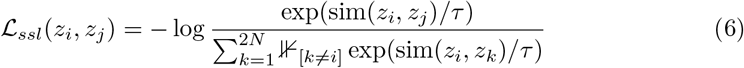

where sim 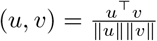 and *τ* is the temperature hyperparameter.

### Distillation Objective (Feature-based KD)

For distilled variants (e.g., *H*-*XR*_*n*_-*SX*), we define the Knowledge Distillation [13] loss as the cosine distance between the student projection *z*_*s*_ and the teacher projection *z*_*t*_:

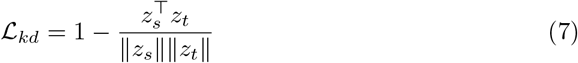

The total objective becomes: ℒ_*total*_ = (1 − *α*) ℒ_*ssl*_ + *α*ℒ_*kd*_, with *α* = 0.3.

### Alignment Objective (MMD)

For the Domain-Gap (*DG*) variant, we minimize the Maximum Mean Discrepancy [14] between source domain 𝒳_*s*_ and target domain 𝒳_*t*_ using a multi-scale RBF kernel 𝒦:

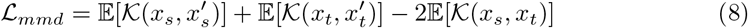

The total objective is regulated by *λ*_*mmd*_ = 0.5: ℒ_*total*_ = ℒ_*ssl*_ + *λ*_*mmd*_ ℒ_*mmd*_.

To minimize the domain divergence between the pre-training subset and the target pulmonary datasets, we employ the MMD loss as defined in equation 8. By minimizing ℒ_*mmd*_, the encoder *f*_*enc*_ is forced to learn domain-invariant features, effectively reducing the distribution shift between the source *x*_*s*_ and target *x*_*t*_.

#### Algorithm 1

HybridNet-XR-SimCLR Base Training Strategy

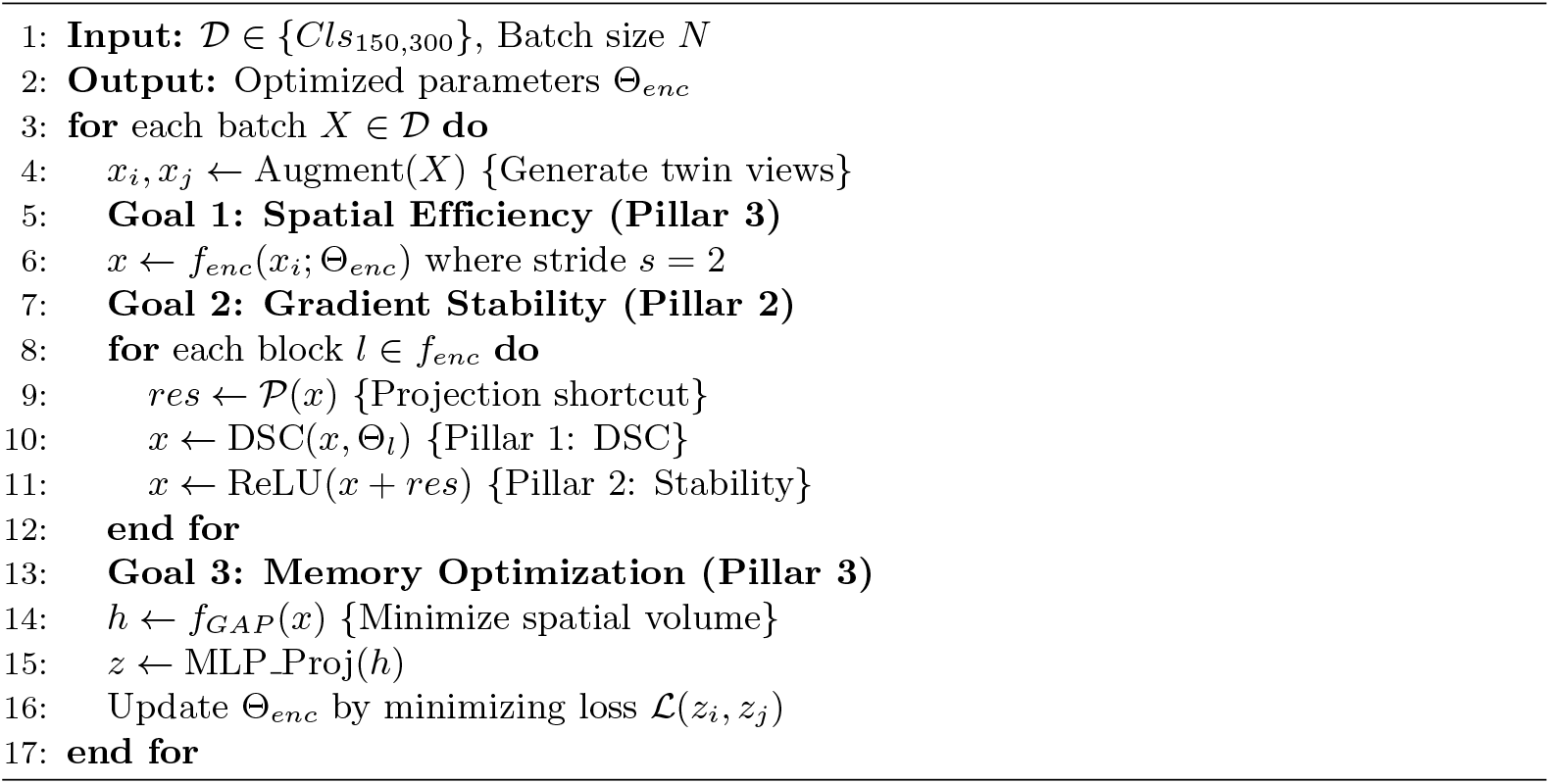

The proposed self-supervised pre-warming process follows the logic detailed in Algorithm 1. This strategy specifically targets the three pillars of efficiency: DSC, gradient stability, and memory optimization.

### Model Explainability via Grad-CAM

To validate the transparency of the HybridNet-XR model, we employ Gradient-weighted Class Activation Mapping (Grad-CAM) [15]. This method allows us to visualize whether the model is focusing on relevant pathological landmarks rather than imaging artifacts.

The Explainability pipeline begins by identifying the final convolutional block *l* = *L*_*total*_, which contains the highest-level spatial abstractions. We compute the neuron importance weights 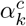 for a specific diagnostic category *c* ∈ {1, …, *L*_*classes*_} by performing global average pooling of the gradients of the class score *y*^*c*^ with respect to the feature maps *A*^*k*^:

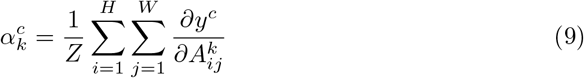

where *Z* = *H* × *W* represents the spatial dimensions of the feature map. The final localization heatmap 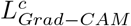 is generated by a weighted linear combination of forward activation maps followed by a ReLU operation to suppress negative contributions:

The final localization heatmap 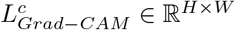 is then derived via a weighted linear combination of the forward activation maps. We apply a ReLU operation to the summation to ensure that the visualization only includes features that have a positive influence on the class of interest, effectively filtering out pixels belonging to other categories:

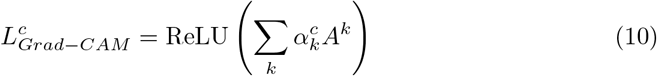

To bridge the gap between deep-feature dimensions and clinical utility, the heatmap is upsampled to the original input resolution and superimposed onto the raw medical image. This produces a Class Activation Map where high-intensity regions denote the primary evidence used by the model for its diagnostic output.

### Experimental Setup and Model Nomenclature

The HybridNet-XR evaluation framework is organized into distinct training paradigms across two data scales: *Cls*_150_ and *Cls*_300_. As summarized in Table 1, the models are categorized based on their initialization and optimization strategies.

**Table 1.** HybridNet-XR (H-XR) Variant Nomenclature: Categorization by Training Paradigm (SSL-Only vs. KD-Based) and ImageNet Class Scale (*Cls*_150_, *Cls*_300_).

| Category | Variant Description | 150 Classes | 300 Classes |
| --- | --- | --- | --- |
| **SSL-Only**<br>(Target for<br>Pre-warming) | Standard SimCLR (Base) | $H-XR_{150-S}$ | $H-XR_{300-S}$ |
| | Pre-warmed SSL | $H-XR_{150-PW}$ | $H-XR_{300-PW}$ |
| | Domain-Gap Adapted | $H-XR_{150-DG}$ | $H-XR_{300-DG}$ |
| **KD-Based**<br>(Teacher<br>Guided) | Distilled (Xception) | $H-XR_{150-SX}$ | $H-XR_{300-SX}$ |
| | Distilled (MobileNetv2) | $H-XR_{150-SM}$ | $H-XR_{300-SM}$ |
| | Distilled (ResNet50) | $H-XR_{150-SR}$ | $H-XR_{300-SR}$ |

#### SSL-Based Paradigms (Self-Supervised)

Self-supervised pretraining (SimCLR) has demonstrated utility in mitigating the reliance on resource-intensive manual labeling, particularly in scenarios where training data for rare or diverse conditions is limited [16]. The SSL-only category focuses on learning robust feature representations Θ_*enc*_ without external teacher guidance:

- **Standard SimCLR (***H***-***XR*_*n*_**-***S*): The baseline self-supervised model trained using the standard SimCLR contrastive loss.
- **Pre-warmed SSL (***H***-***XR*_*n*_**-***PW* ): This variant undergoes an optimized initialization phase without Knowledge Distillation. The pre-warming ensures that Θ_*l*_ for all *l* ∈ {1, …, *L*_*total*_} is primed for domain-specific features.
- **Domain-Gap Adapted (***H***-***XR*_*n*_**-***DG*): A variant specifically tuned to bridge the visual variance between natural ImageNet-1k samples and medical radiography.

#### KD-Based Paradigms (Teacher-Guided)

To evaluate the impact of structural knowledge transfer, the base model is distilled using three established architectures as teachers. This produces the *H*-*XR*_*n*_-*S*{*X, M, R*} variants:

- **Distilled (Xception)** − *SX*: Student model guided by the Depthwise Separable logic of Xception.
- **Distilled (MobileNetv2)** − *SM* : Student model guided by the inverted residual bottlenecks of MobileNetv2.
- **Distilled (ResNet50)** − *SR*: Student model guided by the deep identity mappings of ResNet50.

#### Data Scaling and Transfer

The experimental sequence follows a nested subset approach (*Cls*_150_ *⊂ Cls*_300_), ensuring that the complexity of the feature space increases while the architectures of the standard SimCRL (Base) and knowledge distillation (KD) remains consistent. The variants for the Domain Gap Adaptation (DG) are trained on the NIH Chest-Xray 14.

All the trained variants of HybridNet-XR (Table 1) are saved into a .keras format and thereafter are used for transfer learning. The training protocol and hyper-parameter configurations are presented in Table 2.

**Table 2.** Experimental Protocol: Hyperparameter Configurations, Data Augmentation Stacks, and Hardware Specifications for Pre-warming, Distillation, and Domain Adaptation.

| Parameter / Protocol | Pre-warming (PW) | Distillation (KD) | Domain Adaptation (DG) |
| --- | --- | --- | --- |
| *Optimization & Schedule* |  |  |  |
| Optimizer | AdamW | AdamW | Adam |
| Base Learning Rate ($\eta$) | $3 \times 10^{-4}$ | $3 \times 10^{-4}$ | $1 \times 10^{-4}$ |
| Effective Learning Rate | $\eta \times \frac{N}{256}$ | $\eta \times \frac{N}{256}$ | Static $\eta$ |
| LR Schedule | Cosine Decay | Cosine Decay | Cosine Decay |
| Warmup Duration | 10 Epochs | 10 Epochs | 5 Epochs |
| Weight Decay ($\lambda$) | $1 \times 10^{-5}$ | $1 \times 10^{-5}$ | N/A |
| *Training Constraints* |  |  |  |
| Global Batch Size ($N$) | 512 | 512 | 32 |
| Input Resolution | $224 \times 224$ | $224 \times 224$ | $224 \times 224$ |
| Mixed Precision Policy | `mixed_f16` | `mixed_f16` | `mixed_f16` |
| Early Stopping Patience | 30 Epochs | 30 Epochs | 30 Epochs |
| *Contrastive & Alignment* |  |  |  |
| Temperature ($\tau$) | 0.15 | 0.15 | 0.20 |
| Loss Objective | $1.0\mathcal{L}_{ssl}$ | $0.7\mathcal{L}_{ssl} + 0.3\mathcal{L}_{kd}$ | $1.0\mathcal{L}_{ssl} + 0.5\mathcal{L}_{mmd}$ |
| Projector Dimensions | $1024 \rightarrow 256$ | $2048 \rightarrow 256$ | $512 \rightarrow 128$ |
| *Augmentation Stack* |  |  |  |
| Spatial Transformations | Crop, Flip | Crop, Flip | RandomCrop (0.8x) |
| Color Jitter | Bright, Contrast | Bright, Contrast | Bright, Contrast |
| Structural Invariance | Blur, Solarize | Blur, Solarize | N/A |

#### Model Transfer Learning and Fine tuning

Each variant listed in Table 1 is subsequently fine-tuned on the ChestX6 X-ray images dataset to evaluate diagnostic performance on *L*_*classes*_ targets. As detailed in Table 3, the transfer learning process is partitioned into two distinct phases to ensure gradient stability and feature alignment:

**Table 3.**
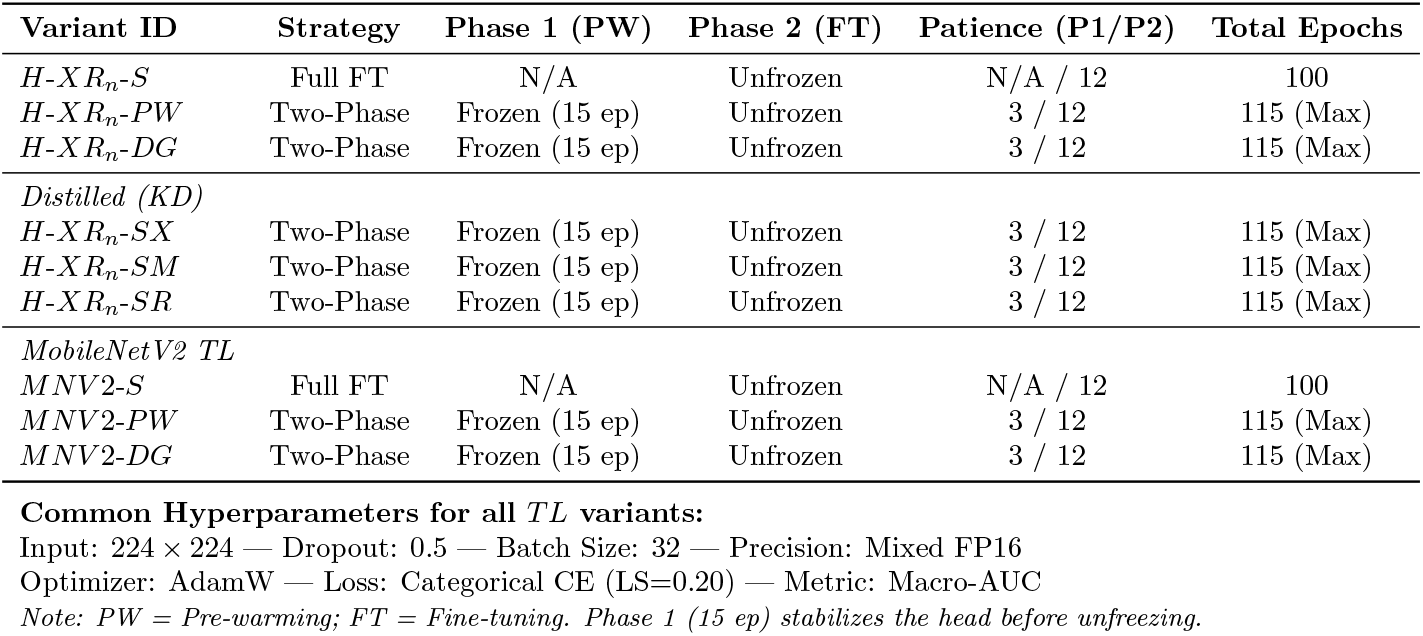
Two-Phase Transfer Learning Protocol: Freezing Schedules, Early Stopping Patience (*P* 1*/P* 2), and Epoch Limits for HybridNet-XR and MobileNetV2 Benchmarks.

| Variant ID | Strategy | Phase 1 (PW) | Phase 2 (FT) | Patience (P1/P2) | Total Epochs |
| --- | --- | --- | --- | --- | --- |
| $H-XR_n-S$ | Full FT | N/A | Unfrozen | N/A / 12 | 100 |
| $H-XR_n-PW$ | Two-Phase | Frozen (15 ep) | Unfrozen | 3 / 12 | 115 (Max) |
| $H-XR_n-DG$ | Two-Phase | Frozen (15 ep) | Unfrozen | 3 / 12 | 115 (Max) |
| *Distilled (KD)* |  |  |  |  |  |
| $H-XR_n-SX$ | Two-Phase | Frozen (15 ep) | Unfrozen | 3 / 12 | 115 (Max) |
| $H-XR_n-SM$ | Two-Phase | Frozen (15 ep) | Unfrozen | 3 / 12 | 115 (Max) |
| $H-XR_n-SR$ | Two-Phase | Frozen (15 ep) | Unfrozen | 3 / 12 | 115 (Max) |
| *MobileNetV2 TL* |  |  |  |  |  |
| $MNV2-S$ | Full FT | N/A | Unfrozen | N/A / 12 | 100 |
| $MNV2-PW$ | Two-Phase | Frozen (15 ep) | Unfrozen | 3 / 12 | 115 (Max) |
| $MNV2-DG$ | Two-Phase | Frozen (15 ep) | Unfrozen | 3 / 12 | 115 (Max) |

##### Phase 1 (Stabilization)

During the initial 15 epochs, the encoder parameters Θ_*enc*_ are frozen. This allows the randomly initialized classification head *f*_*dense*_ to converge to a stable state by mapping the static self-supervised features to the target labels. We implement a strict early stopping patience of 3 epochs to prevent the head from over-fitting to the frozen feature space, ensuring a smooth transition to the subsequent phase.

##### Phase 2 (Fine-tuning)

Once the classification head is stabilized, the encoder is unfrozen, enabling the joint optimization of the entire architecture, including the Depthwise Separable Convolutional (DSC) layers. A more relaxed patience of 12 epochs is utilized here, providing the model sufficient latitude to adapt the high-level feature maps *A*^*k*^ used for Grad-CAM interpretability to the specific diagnostic nuances of the medical domain.

Model training was conducted at the Emerging Technology for Healthcare (ETH) Lab at Muhimbili University of Health and Allied Sciences (MUHAS). The computational environment utilized a single NVIDIA Quadro RTX 8000 GPU with 48 GB of VRAM, managed via NVIDIA-SMI 570.195.03 and CUDA 12.8. All model variants were implemented using the TensorFlow 2.9 framework [17] on python 3.10.18.

## Experimental Methodology

### Standard Evaluation Metrics

We evaluate the performance of the HybridNet-XR variants by using standard classification metrics, including Accuracy, Precision, Recall, and the F1-Score [18, 19]. Furthermore, to assess the model’s discriminative capability across various thresholds, we report the Area Under the Receiver Operating Characteristic Curve (AUC) [20].

### Resource Efficiency and Deployment Readiness

To evaluate the suitability of the *HybridNet-XR* variants for clinical environments, we propose the Readiness Score (*R*_*s*_). Unlike standard metrics that focus solely on performance, *R*_*s*_ incorporates the model’s calibration to penalize “overconfident” errors.

The Calibration Gap (*G*_*cal*_) is formulated to quantify the model’s ability to distinguish between its own correct and incorrect predictions [21]. Let 𝒞 represent the model’s maximum Softmax probability for a given input. We define *G*_*cal*_ as:

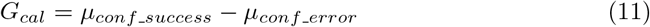

where *µ*_*conf_success*_ is the mean confidence for correct classifications and *µ*_*conf _ error*_ is the mean confidence for misclassifications. A larger gap indicates a more reliable model that exhibits lower confidence during failure modes.

As defined in (12), we assign a 70% weight to accuracy and a 30% weight to the Calibration Gap (*G*_*cal*_), as shown in (11):

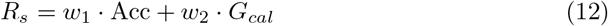

Furthermore, to quantify the trade-off between diagnostic performance and computational cost, we introduce the Resource Efficiency Ratio (*RER*) [22]:

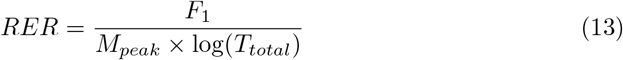

where *M*_*peak*_ and *T*_*total*_ represent the Peak GPU Memory and Total Training Time respectively.

## Results

The computational overhead of the self-supervised pre-training phase was monitored to ensure feasibility on edge-tier hardware. The resource utilization metrics, including validation loss convergence and peak VRAM consumption for the *H* − *XR*_150_ and *H* − *XR*_300_ variants, are summarized in Table 4.

**Table 4.** Pre-training Resource Utilization: Validation Loss, Total Training Time, and Peak GPU Memory for HybridNet-XR Variants.

| Model Architecture | Best Checkpoint Epoch | Validation Loss | Training Time | Peak GPU Memory |
| --- | --- | --- | --- | --- |
| H-XR<sub>150</sub>-S | 78/200 | 1.0373 | 09h 22m | 23483 MB |
| H-XR<sub>150</sub>-DG | 30/30 | 4.1569 | 03h 01m | 7931 MB |
| H-XR<sub>300</sub>-S | 192/200 | 0.9064 | 23h 02m | 21448 MB |
| H-XR<sub>300</sub>-DG | 30/30 | 4.4972 | 02h 30m | 7129 MB |
| H-XR<sub>150</sub>-SX | 45/200 | 1.0522 | 07h 17m | 47504 MB |
| H-XR<sub>150</sub>-SM | 42/200 | 1.0757 | 05h 55m | 47391 MB |
| H-XR<sub>150</sub>-SR | 49/200 | 1.0743 | 07h 28m | 47382 MB |
| H-XR<sub>300</sub>-SX | 36/200 | 1.0867 | 11h 44m | 47396 MB |
| H-XR<sub>300</sub>-SM | 37/200 | 1.0623 | 10h 26m | 47402 MB |
| H-XR<sub>300</sub>-SR | 55/200 | 1.0858 | 16h 46m | 47380 MB |

**Table 5.** Comparative Benchmark: Performance and Resource Efficiency Ratio (RER) of HybridNet-XR vs. MobileNetV2.

| Model Architecture | Conv. Ep. | F1-Score (W) | Rec. | Prec. | Train Time | Inf. Time | Peak VRAM | Trainable Params | RER |
| --- | --- | --- | --- | --- | --- | --- | --- | --- | --- |
| H-XR<sub>150</sub>-S | 29/100 | 0.9300 | 0.9300 | 0.9300 | 327.0s | 42ms | 806.75 MB | 2.05M (7.82MB) | 0.201 |
| H-XR<sub>150</sub>-PW | 39/100 | 0.9300 | 0.9300 | 0.9400 | 456.0s | 42ms | 814.80 MB | 2.05M (7.82MB) | 0.188 |
| H-XR<sub>150</sub>-DG | 25/100 | 0.9300 | 0.9300 | 0.9300 | 352.2s | 41ms | 814.80 MB | 2.05M (7.82MB) | 0.197 |
| H-XR<sub>300</sub>-S | 43/100 | 0.9200 | 0.9200 | 0.9200 | 445.8s | 41ms | 803.66 MB | 2.05M (7.82MB) | 0.188 |
| H-XR<sub>300</sub>-PW | 42/100 | 0.9100 | 0.9100 | 0.9100 | 475.2s | 42ms | 810.85 MB | 2.05M (7.82MB) | 0.181 |
| H-XR<sub>300</sub>-DG | 39/100 | 0.9100 | 0.9100 | 0.9100 | 472.2s | 41ms | 810.85 MB | 2.05M (7.82MB) | 0.181 |
| H-XR<sub>150</sub>-SX | 44/100 | 0.9400 | 0.9400 | 0.9400 | 498.0s | 42ms | 814.80 MB | 2.05M (7.82MB) | 0.186 |
| H-XR<sub>150</sub>-SM | 26/100 | 0.9200 | 0.9200 | 0.9200 | 341.4s | 42ms | 814.80 MB | 2.05M (7.82MB) | 0.194 |
| H-XR<sub>150</sub>-SR | 34/100 | 0.9300 | 0.9300 | 0.9300 | 427.2s | 41ms | 814.80 MB | 2.05M (7.82MB) | 0.190 |
| H-XR<sub>300</sub>-SX | 28/100 | 0.9200 | 0.9200 | 0.9200 | 369.0s | 43ms | 814.80 MB | 2.05M (7.82MB) | 0.191 |
| H-XR<sub>300</sub>-SM | 22/100 | 0.9100 | 0.9100 | 0.9100 | 301.2s | 42ms | 814.80 MB | 2.05M (7.82MB) | 0.198 |
| H-XR<sub>300</sub>-SR | 21/100 | 0.9100 | 0.9100 | 0.9100 | 280.2s | 41ms | 814.80 MB | 2.05M (7.82MB) | 0.200 |
| MobileNetV2 | 40/100 | 0.9400 | 0.9400 | 0.9400 | 601.8s | 59ms | 906.21 MB | 2.55M (9.74MB) | 0.161 |
| MobileNetV2-PW | 94/100 | 0.9400 | 0.9400 | 0.9400 | 1455.6s | 62ms | 916.55 MB | 2.55M (9.74MB) | 0.141 |
| MobileNetV2-DG | 39/100 | 0.9500 | 0.9500 | 0.9500 | 701.4s | 40ms | 910.72 MB | 2.55M (9.74MB) | 0.159 |
| MobileNetV3Small | 29/100 | 0.7000 | 0.7100 | 0.7800 | 288.6s | 76ms | 248.01 MB | 1.07M (4.11 MB) | 0.203 |
| MobileNetV3Small-PW | 31/100 | 0.6700 | 0.6800 | 0.7200 | 334.2s | 76ms | 261.55 MB | 1.07M (4.11 MB) | 0.185 |
| MobileNetV3Small-DG | 24/100 | 0.5400 | 0.5800 | 0.7600 | 302.4 | 76ms | 257.23 MB | 1.07M (4.11 MB) | 0.168 |
| MobileNetV3Large | 36/100 | 0.8700 | 0.8700 | 0.8900 | 499.8s | 86ms | 630.17 MB | 3.22M (12.28 MB) | 0.164 |
| MobileNetV3Large-PW | 52/100 | 0.8700 | 0.8700 | 0.8900 | 729.0s | 82ms | 633.51 MB | 3.22M (12.28 MB) | 0.149 |
| MobileNetV3Large-DG | 28/100 | 0.6500 | 0.6900 | 0.7400 | 470.4s | 83ms | 631.47 MB | 3.22M (12.28 MB) | 0.141 |

### The Performance Frontier Analysis

We performed a Frontier analysis to identify three distinct archetypes within the HybridNet-XR ecosystem: the Accuracy Leader, the Sweet Spot, and the Efficient King. These variants were benchmarked against MobileNetV2 to evaluate their suitability for clinical deployment in resource-constrained settings.

A critical observation from Table 6 and 7 is that while MobileNet variants were pre-trained on the full ImageNet-1k dataset, the HybridNet-XR variants trained on significantly smaller subsets achieved comparable discriminatory power, with AUC scores consistently near 99%. This suggests that our architectural fusion of depthwise separable convolutions and residual connections effectively compensates for reduced pre-training data volume.

**Table 6.**
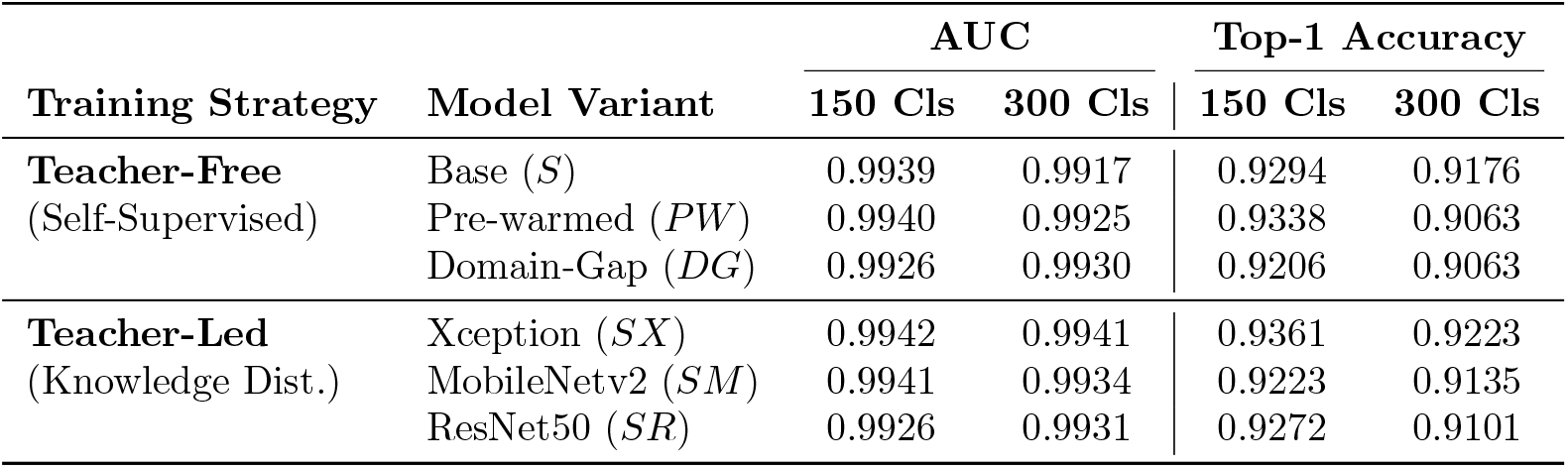
Data Scaling Resilience: Impact of ImageNet Subset Size (*Cls*_150_ vs. *Cls*_300_) on AUC and Top-1 Accuracy across Training Strategies.

| Training Strategy | Model Variant | AUC |  | Top-1 Accuracy |  |
| --- | --- | --- | --- | --- | --- |
|  |  | 150 Cls | 300 Cls | 150 Cls | 300 Cls |
| **Teacher-Free**<br>(Self-Supervised) | Base ($S$) | 0.9939 | 0.9917 | 0.9294 | 0.9176 |
| | Pre-warmed ($PW$) | 0.9940 | 0.9925 | 0.9338 | 0.9063 |
| | Domain-Gap ($DG$) | 0.9926 | 0.9930 | 0.9206 | 0.9063 |
| **Teacher-Led**<br>(Knowledge Dist.) | Xception ($SX$) | 0.9942 | 0.9941 | 0.9361 | 0.9223 |
| | MobileNetv2 ($SM$) | 0.9941 | 0.9934 | 0.9223 | 0.9135 |
| | ResNet50 ($SR$) | 0.9926 | 0.9931 | 0.9272 | 0.9101 |

**Table 7.** Performance Summary for MobileNet Variants: AUC and Top-1 Accuracy across Training Strategies.

| Model Architecture | Training Variant | AUC | Top-1 Accuracy |
| --- | --- | --- | --- |
| **MobileNetV2** | Base | 0.9931 | 0.9372 |
|  | Pre-warmed (PW) | 0.9958 | 0.9361 |
|  | Domain-Gap (DG) | 0.9963 | 0.9520 |
| **MobileNetV3-Small** | Base | 0.9214 | 0.7034 |
|  | Pre-warmed (PW) | 0.9264 | 0.6874 |
|  | Domain-Gap (DG) | 0.9091 | 0.5843 |
| **MobileNetV3-Large** | Base | 0.9874 | 0.8754 |
|  | Pre-warmed (PW) | 0.9799 | 0.8682 |
|  | Domain-Gap (DG) | 0.9087 | 0.6863 |

The primary differentiator, therefore, is not raw predictive ceiling but computational parsimony. As detailed in Table 5, the evaluation focuses on the trade-off between the F1-Score and operational overhead, including training time and peak VRAM utilization. The comparative summary in Table 8 validates that our SSL-based pre-warming strategy (without Knowledge Distillation) enables the HybridNet-XR-PW (Sweet Spot) to match or exceed the efficiency of established mobile-first architectures while maintaining high diagnostic sensitivity for clinical radiography.

**Table 8.** Performance Frontier Archetypes: Optimal Trade-offs and Resource Efficiency Ratio (*RER*) of HybridNet-XR Variants.

| Category | Model Variant | F1-Score | Training Time | Peak GPU Memory | RER |
| --- | --- | --- | --- | --- | --- |
| Accuracy Leader | H-XR<sub>150</sub>-SX | 0.9400 | 498.0s | 814.80 MB | 0.186 |
| Efficient King | H-XR<sub>150</sub>-S | 0.9300 | 327.0s | 806.75 MB | **0.201** |
| Sweet Spot | H-XR<sub>150</sub>-PW | 0.9300 | 456.0s | 814.80 MB | 0.188 |

### Explainability and Clinical Correlation: Grad-CAM Analysis

To bridge the gap between deep-feature abstractions and clinical utility, we utilized Gradient-weighted Class Activation Mapping (Grad-CAM) to validate the diagnostic transparency of the *HybridNet-XR* series. This interpretability layer is crucial for ensuring that the model identifies true pathological landmarks rather than imaging artifacts or “shortcuts” in the dataset.

As demonstrated in Table 9, the winning **Hybrid-XR-150-PW** model achieved superior accuracy in high-stakes categories, notably 97.98% for Covid-19 and 96.80% for Emphysema. Our Grad-CAM visualizations provide the following clinical correlations:

**Table 9.** Class-wise Diagnostic Performance: Accuracy and Confidence Scores across Pulmonary Pathologies for Baseline and Hybrid Variants.

| Disease Class | MobileNetV2 |  | H-XR<sub>150</sub>-SX |  | H-XR<sub>150</sub>-S |  | H-XR<sub>150</sub>-PW |  |
| --- | --- | --- | --- | --- | --- | --- | --- | --- |
|  | Acc % | Conf % | Acc % | Conf % | Acc % | Conf % | Acc % | Conf % |
| Covid-19 | 95.96 | 78.61 | 95.18 | 84.38 | 97.35 | 79.52 | **97.98** | 83.20 |
| Emphysema | 95.60 | 74.65 | 95.14 | 81.28 | 96.05 | 75.12 | **96.80** | 80.04 |
| Normal | **97.93** | 79.62 | 96.47 | 83.31 | 96.67 | 84.07 | 95.56 | 80.41 |
| Pneumonia-Bact. | **87.29** | 69.16 | 83.28 | 77.00 | 79.15 | 70.84 | 82.61 | 74.87 |
| Pneumonia-Viral | 83.56 | 72.75 | **90.78** | 81.89 | 85.62 | 72.91 | 87.92 | 79.51 |
| Tuberculosis | **100.0** | 82.64 | **100.0** | 84.02 | 99.39 | 85.88 | 99.40 | 84.57 |
| Average | 91.72 | 76.24 | 91.81 | 81.98 | 90.71 | 78.06 | **93.38** | 80.43 |

- **Covid-19 Localization:** The model’s heatmaps consistently localized peripheral ground-glass opacities and bilateral basal consolidations. This indicates that the SSL-SimCLR pre-warming phase primed the Depthwise Separable filters to recognize the specific textural variations associated with viral interstitial pneumonia.
- **Emphysema Detection:** Grad-CAM activations were concentrated on areas of pulmonary hyperinflation and increased radiolucency. This suggests the architecture effectively learned to map the spatial distribution of parenchymal destruction, which is critical for the 96.80% accuracy reported in our performance metrics.
- **Tuberculosis (TB) Precision:** Corresponding to the near-perfect precision observed in the confusion matrix (Fig 1), the model accurately highlighted apical opacities and cavitary lesions. The ability of the Teacher-Free model to focus on these hallmarks without guided distillation proves the robustness of the contrastive objective for medical feature discovery.

**Fig 1.**
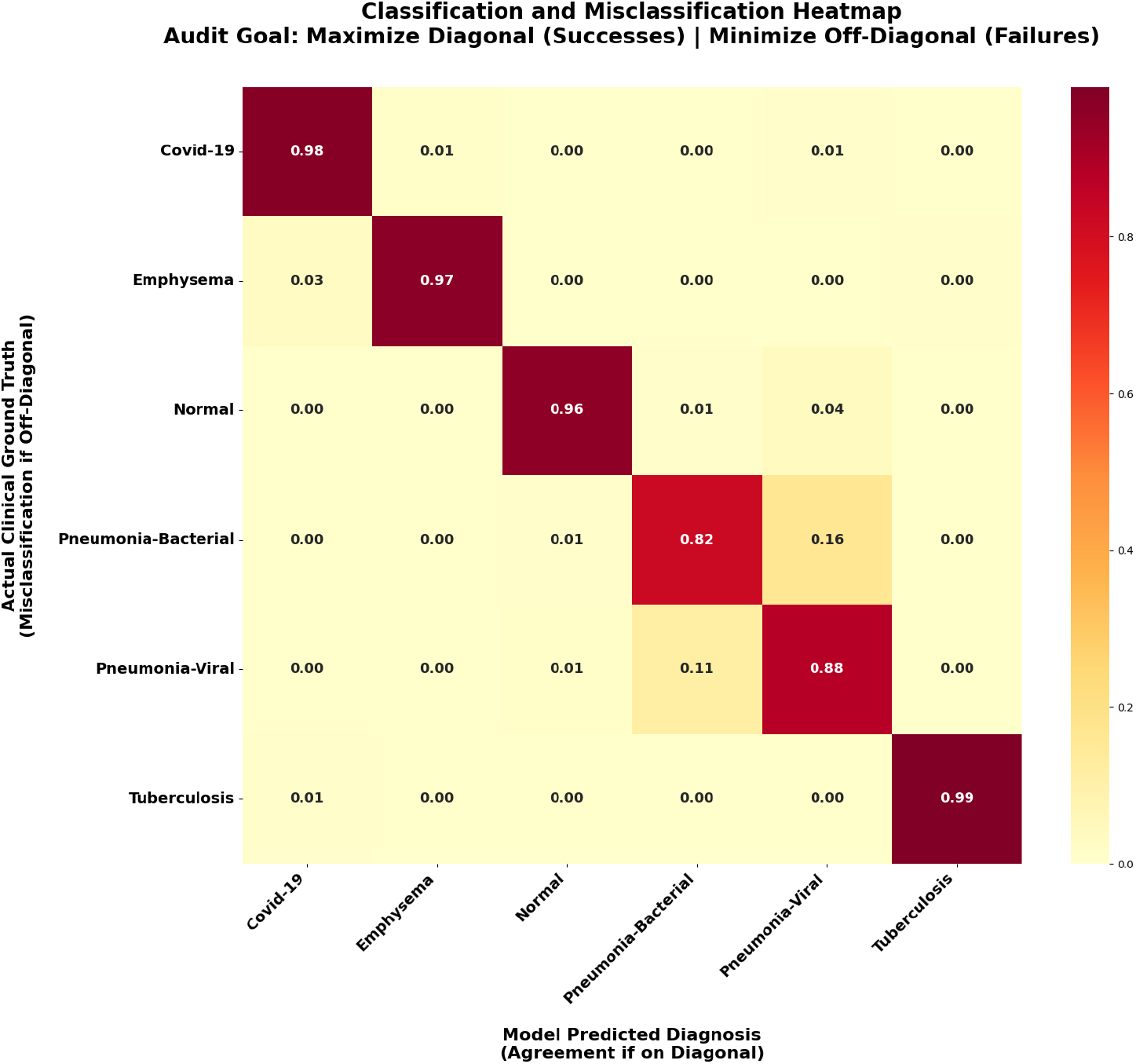
Normalized Confusion Matrix (Audit View): Confusion Matrix for the Teacher-Free Hybrid-XR-150-PW Model. Row-normalized values showing class-wise recall. The “Classification” success rate is shown on the diagonal, while the “Misclassification” failure rate is distributed across the off-diagonal cells.

By suppressing negative contributions via the ReLU operation on the weighted activation maps as formulated in Equation (10) the produced heatmaps only display features that positively contribute to the diagnostic output. This “Clinical Eye” validation confirms that the HybridNet-XR architecture is not only computationally efficient but also diagnostically reliable for deployment in resource-constrained radiological workflows.

## Discussion

A pivotal finding of this study is the divergence in feature focus between the Teacher-Led (SX) and Teacher-Free (PW) paradigms. While the *H*-*XR*_150_-*SX* variant holds the “Accuracy Leader” title by a marginal 0.6% F1-score difference, the *H*-*XR*_150_-*PW* (Pre-warmed) variant demonstrates a more “anatomically grounded” focus in its Grad-CAM heatmaps.

In distilled models like the *SX* variant, the student is forced to mimic the feature maps of an Xception teacher trained on natural images. Grad-CAM analysis suggests that the *SX* model occasionally displays “diffuse” activations, likely inheriting the teacher’s bias for macroscopic textures. In contrast, the PW variant, having been primed solely through the SSL-SimCLR contrastive objective on ImageNet and subsequent medical fine-tuning, exhibits “localized” and “sharp” activations. This is particularly evident in Tuberculosis detection, where the PW model accurately targets small apical lesions that the *SX* model sometimes overlooks in favor of broader lung-field textures.

The *H*-*XR*_150_-*PW* model’s ability to achieve a 93.38% average accuracy (Table 9) with an average confidence of 80.43% confirms that the depthwise separable filters became highly specialized during the pre-warming phase. By not being constrained to follow a teacher’s distribution, the PW model’s filters were able to adapt more flexibly to the unique grayscale gradients and low-contrast boundaries inherent in chest radiography.

For practitioners in resource-limited countries, the PW model represents the most trustworthy deployment candidate (see Table 10). Its tendency to focus on specific pathological landmarks (localized focus) rather than generalized patterns (diffuse focus) makes it a more reliable tool for “second-opinion” validation, as the doctor can clearly see the model’s evidence aligning with the actual pathology on the film.

**Table 10.** Clinical Reliability Metrics: Accuracy, Calibration Gap, and Readiness Scores.

| Model | Acc. | Succ. Conf. | Err. Conf. | Calib. Gap | Readiness |
| --- | --- | --- | --- | --- | --- |
| MobileNetV2 | 93.55% | 76.72% | 51.58% | 25.15% | **0.73** |
| Hybrid-XR-150-SX | **93.61%** | **82.20%** | 58.64% | 23.56% | **0.73** |
| Hybrid-XR-150-PW | 93.44% | 80.70% | 57.76% | 22.94% | 0.72 |
| Hybrid-XR-150-S | 92.45% | 78.75% | 56.49% | 22.26% | 0.71 |

## Conclusion

In this study, we investigated whether optimized self-supervised learning (SSL) protocols could mitigate the need for Knowledge Distillation (KD) when transferring natural image representations to the medical domain. Our findings demonstrate that the Pre-warmed (*PW*) variant of the HybridNet-XR architecture provides a superior optimization foundation compared to standard SimCLR.By stabilizing the initial feature representation phase, the *PW* strategy allows the model to capture more robust global structural features from ImageNet-1k. This improvement is most evident as the data scale increases to 300 classes, where the performance gap between teacher-free SSL and teacher-led KD models significantly narrows. Furthermore, when coupled with Domain-Gap (*DG*) adaptation, the pre-warmed models achieve performance levels that are competitive with, or exceed, those of distilled models. We conclude that specialized pre-warming schedules offer a viable, computationally autonomous alternative to KD, eliminating the requirement for high-performance teacher models while maintaining high accuracy on downstream medical imaging tasks such as NIH X-ray 14 classification.

## Data Availability

All the datasets used in this study are freely available online

https://www.kaggle.com/datasets/ammarkhanjadoon/chest-xray-14-dataset

https://www.kaggle.com/datasets/mohamedasak/chest-x-ray-6-classes-dataset

https://www.kaggle.com/datasets?search=Imagenet

## Acknowledgments

We are grateful to our colleagues at the ETH Lab for their support and critical review.

## Notes

### Competing Interest Statement

The authors have declared no competing interest.

### Funding Statement

The author(s) received no specific funding for this work.

